# A systematic scoping review of adult obesity policy actions and weight-related services in a region of the United Kingdom using the Behaviour Change Wheel

**DOI:** 10.1101/2025.03.01.25323144

**Authors:** Aoibhin Kelly, Helen Croker, Roisin O’Neill, Jayne V. Woodside, Laura McGowan

## Abstract

Obesity is a global health challenge associated with increased risk of co-morbidity and mortality. Existing government policies have failed to adequately address obesity. Mapping obesity policy actions (including prevention initiatives and weight-management services (WMS)) using behavioural science can offer insights into how approaches to prevention/management could be improved.

This research aimed to identify strengths and gaps in adult obesity policy actions/WMS in Northern Ireland (NI) by conducting a systematic scoping review of grey literature. Obesity policy actions/WMS were mapped out and coded using the Behaviour Change Wheel (BCW), then assessed against obesity risk factors identified from the Foresight Obesity System Map.

The breadth of obesity policy actions/WMS was mapped using Google Advanced Searches, grey literature database searches, targeted website searches and stakeholder consultations. Policy actions/WMS were categorised using the UK weight-management tiered system. Policy actions/WMS were coded by BCW intervention type. Foresight variables were coded according to the Capability-Opportunity-Motivation-Behaviour (COM-B) Model. Policy actions/WMS were subsequently mapped against Foresight variables and displayed using a heat map.

Twelve Google Advanced Searches, three database searches, n=17 targeted website searches and three stakeholder interviews were conducted (and updated in 2024) identifying a wide range of relevant records (N=127). Results included policies/strategies/guidelines/campaigns/services targeting weight management/obesity.

Of the identified policy actions/WMS, 72% were classified as tier 1 (health promotion); 21% tier 2 (community-based weight management programmes); 2% tier 3 (specialist obesity services), 5% were tier 2/3 services and 0% were tier 4 services (metabolic surgery). Education and persuasion were the most commonly coded intervention types from the BCW, followed by enablement and training. Environmental restructuring was limited, as was modelling and incentivisation approaches.

Behavioural mapping of obesity policy actions/WMS is a novel approach with the potential to influence policy/service development. In NI, using this method illustrated policy gaps and significant opportunities to improve WMS provision.

## Introduction

Overweight and obesity are complex health challenges associated with significant morbidity and mortality worldwide (1, 2). Subsequent government policies have failed to reduce obesity levels globally, and there is an urgent need to better understand how obesity policies influence health outcomes (3).

Obesity is caused by a myriad of factors including biological, psychological, behavioural, social, cultural, environmental and economic factors (4). A recent 30 year analysis of obesity policies in England concluded that government strategies were not implemented or evaluated adequately, they tended toward the less interventionist approach and reliance on individual agency was common (3). Individual-focused policies are typically ineffective when it comes to reducing obesity levels and may also widen health inequalities (5).

Whole systems approaches (WSAs) to obesity incorporate interventions across multiple levels of a wider system and have shown some success, for example in Amsterdam (6). Many obesity prevention interventions in WSAs are agento-structural (structural changes that facilitate healthier choices) or structural in nature (bypassing individual choices in relation to healthy behaviours) (7), which may be more likely to reduce inequalities (3, 5). A recent estimate by the Behavioural Insights Team in the United Kingdom (UK), forecast that the implementation of four structural/agento-structural obesity prevention policies (including restrictions on promotion/advertising of unhealthy foods and the Soft Drinks Industry Levy) could result in a net benefit of £76 billion over the next 25 years (Behavioural Insights Team, 2022). These suggested benefits encompass economic and healthcare savings.

Given that between 26-29% of adults across the different regions of the UK are currently living with obesity (defined as a body mass index of 30kg/m² or more), WMS are an important component of WSAs and better distinction between prevention and management pathways are required according to recent recommendations (8). The Obesity Care Pathway in the UK is comprised of four tiers of intervention (figure 1), with tier 1 aimed at prevention and tiers 2-4 used to describe management and treatment options. Coverage and availability of WMS across all tiers varies considerably across the UK, but are particularly lacking in NI, where tiers 3 and 4 (often referred to as specialist obesity services) are not widely available via the NHS (9, 10).

**Figure 1.**
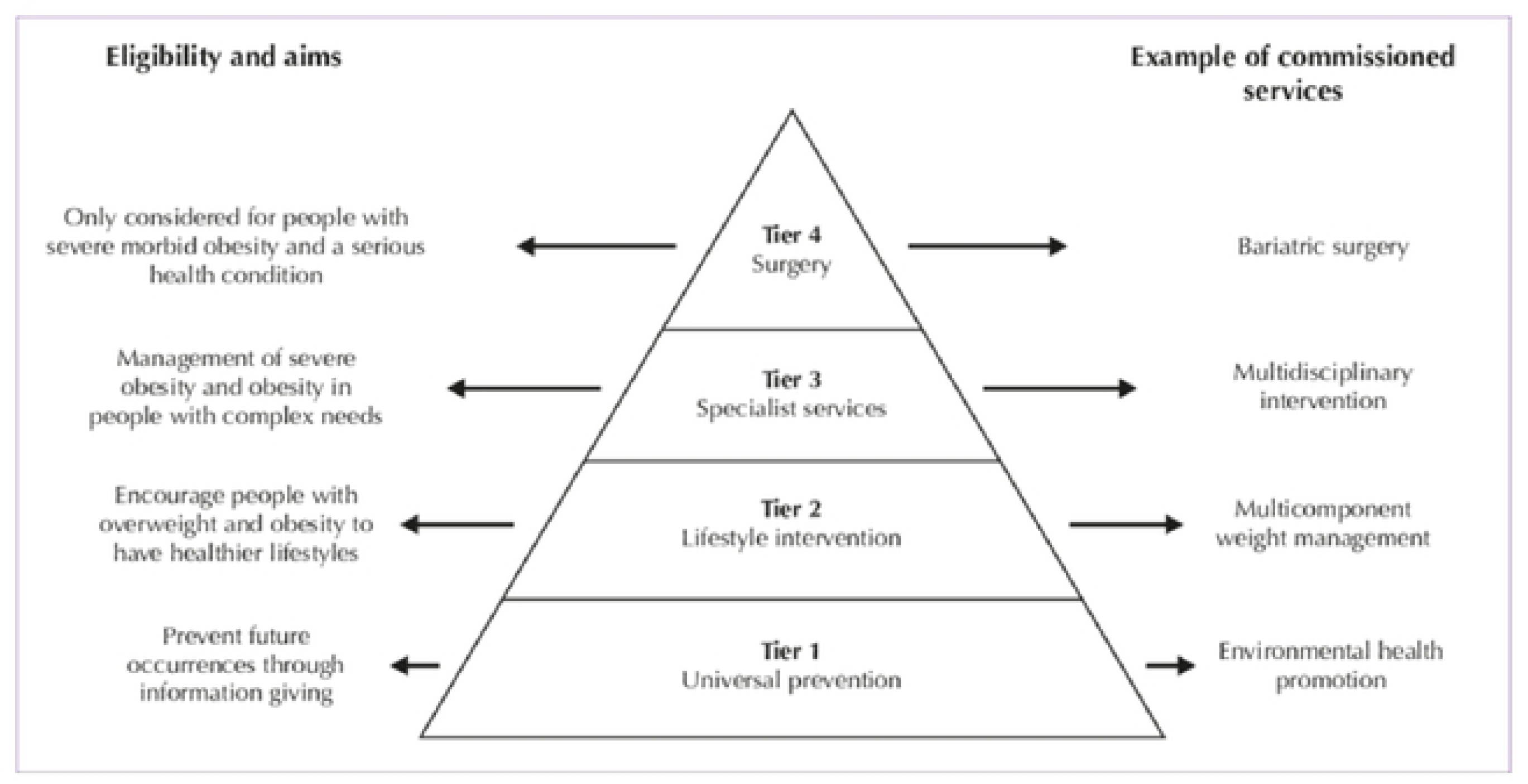
The UK Obesity Care Pathway (Department of Health, 2013)(11)

The NI Assembly Inquiry into Obesity (2009) emphasised the need for dedicated obesity management clinics and a separate bariatric service for those living with obesity, yet no regional obesity care pathway has been implemented. A strategy for addressing overweight and obesity was introduced - “A Fitter Future For All” Framework (2012-2022) however, the targets for reducing population obesity levels were not met. A new obesity strategy for NI, called ‘Healthy Futures’ is in the latter stages of consultation.

Approaches to improve health behaviours can be guided by evidence-based behavioural models, examples of which include the Behaviour Change Wheel (BCW) Framework and the Capability-Opportunity-Motivation-Behaviour (COM-B) Model (12) (see Figure 2). Applying such theoretical approaches should increase the likelihood that policies will successfully change behaviours in the desired manner.

**Figure 2.**
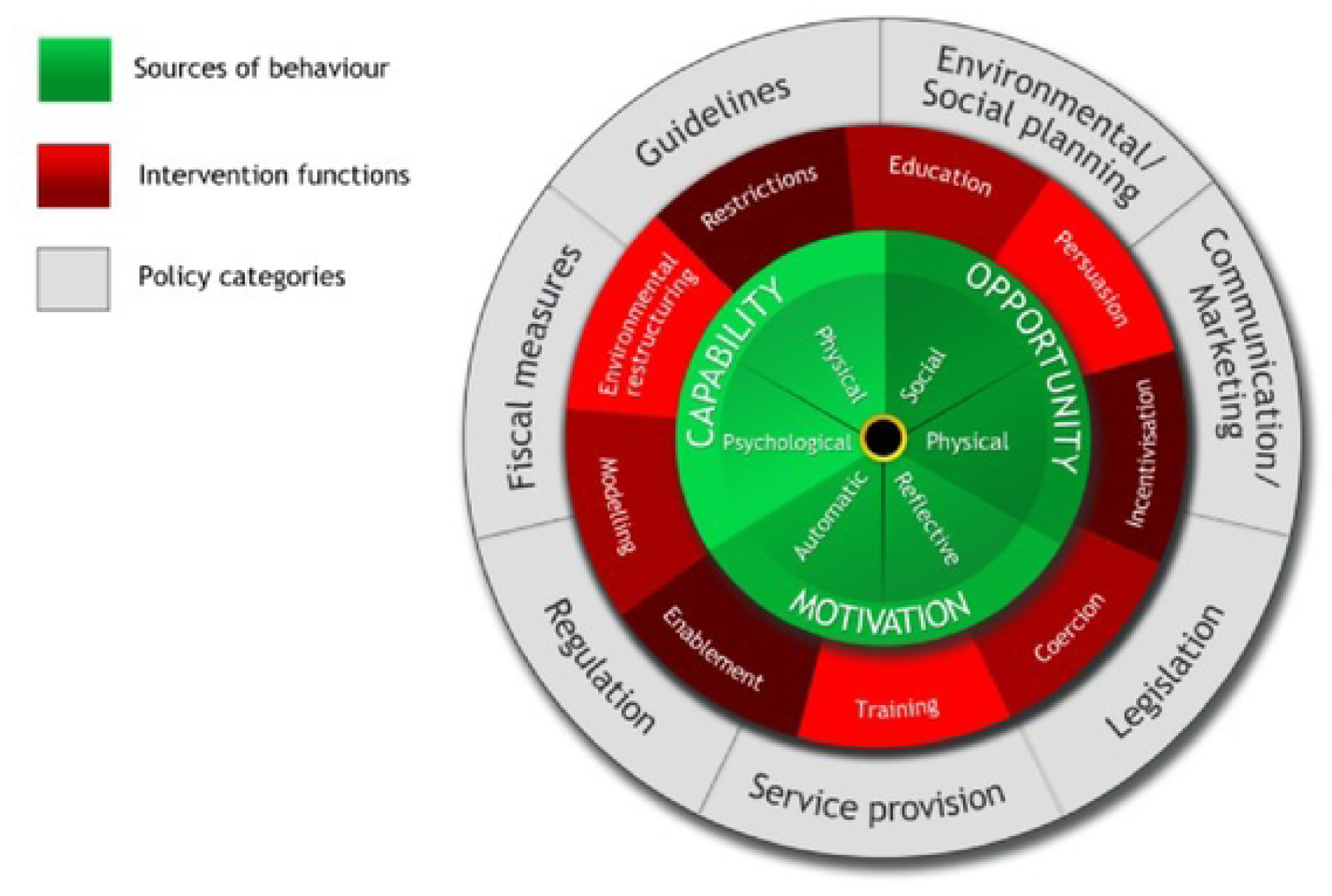
The Behaviour Change Wheel(12)

Previous mapping of early years obesity prevention policies in England, using the BCW and Foresight Obesity System Map, identified gaps in policy. Croker and colleagues selected obesity risk factors from the Foresight Map (coding them by COM-B constructs) and coded policy actions from the literature based on the BCW. Coded policy actions mapped against the key obesity risk factors highlighted areas where obesity prevention efforts were lacking (13). Whilst there was considerable policy activity to address obesity in the early years, efforts were mostly focused on education and guidelines around environmental restructuring (changing the physical or social context) (13). Other studies have demonstrated the utility of policy mapping in health disciplines (14, 15). One study which mapped policy options for early childhood obesity prevention in Australia (and five other countries) found that countries were more likely to have more upstream policies (wider determinants of health e.g., food and built environments) when they had invested in systems-wide approaches e.g., having a national obesity strategy, having separate plans dedicated to food/nutrition and physical activity. However, more often the countries in the study had a higher proportion of downstream/midstream policies. These included clinical/preventive care guidance (downstream) or nutrition promotion guidance/advice to caregivers about childhood obesity (midstream), again demonstrating an emphasis on individual agency (16).

The policy mapping review method by Croker and colleagues (2020) has not been applied to adult obesity, nor have obesity policy actions/WMS been mapped in this way in the UK. Given rising levels of obesity and the complex public health challenges posed by this trend, conducting a systematic scoping review to map existing policies/WMS in NI from a behavioural science perspective has the potential to identify strengths and gaps to guide future obesity policy and service development.

This study’s aims were twofold: 1) to conduct a systematic scoping review which maps obesity policy actions/WMS targeted at adults in NI and categorise them according to UK weight-management tiers; and 2) analyse policy actions/WMS from a behavioural science perspective using the BCW to identify strengths and gaps in provision.

## Methods

A scoping review methodology can identify the breadth of evidence available and map it without critical appraisal, which is suitable for the present research question (17). A systematic grey literature search adapted from Godin and colleagues (18) was conducted to find relevant adult obesity policy actions/WMS. This search methodology involved a 4-step approach to search the grey literature as systematically and robustly as possible. The steps involved, 1) Google Advanced Searches 2) grey literature database searches, 3) targeted website searches, and 4) consultation with contact experts. Examples of search terms and keywords used in the Google Advanced Searches included *∼obesity Northern Ireland AND policies OR services*; and *“weight management” OR “weight loss” AND Northern Ireland* as seen in table 1 of Appendix S1 (full details of the grey literature search methodology including adaptions for the current study also available in Appendix S1). Policy actions/WMS identified were then categorised according to the UK tiered weight-management pathway to map the provision of services across tiers. The Google Advanced Searches and targeted website searches were updated in February 2024.

**Table 1.**
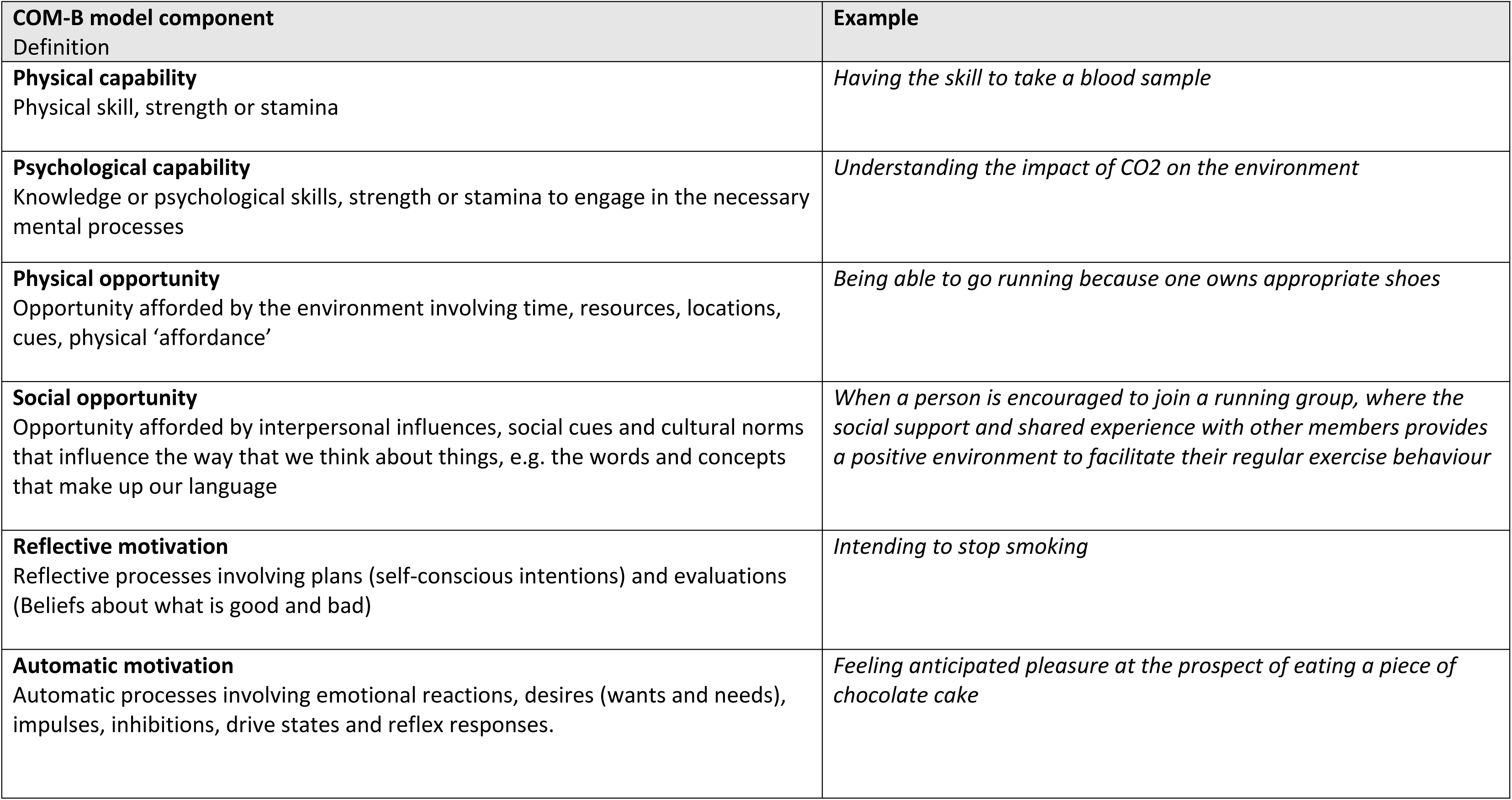
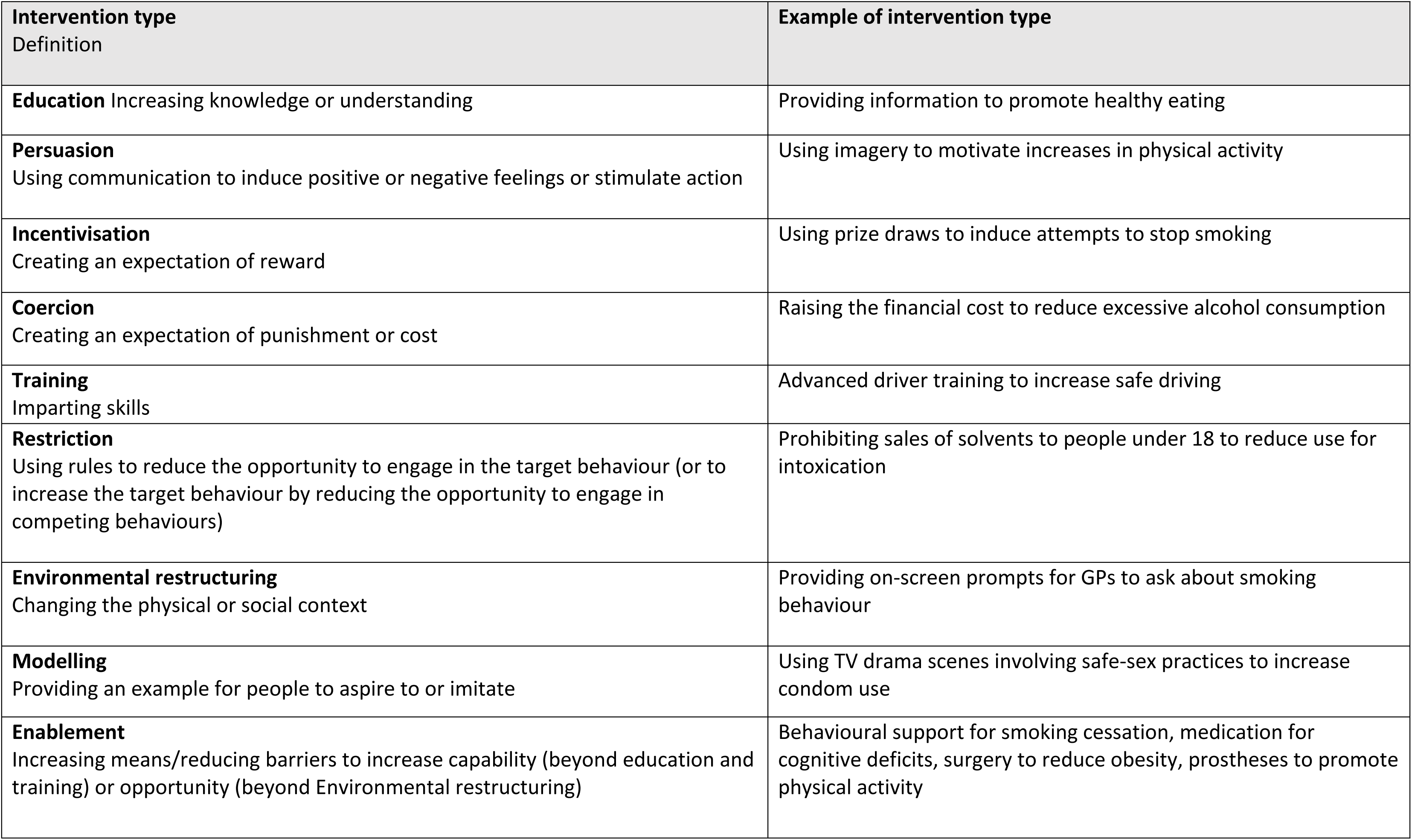
Definitions of COM-B and BCW components (taken from Michie, Atkins and West, 2014, p63, 111,112)

*Inclusion:* Policy actions/WMS included any form of policy, strategy, guideline, campaign, initiative, programme or (NHS-funded) service with a focus on overweight/obesity prevention, adult weight-management/healthy weight and energy-balance behaviours such as healthy eating/increasing physical activity. Childhood obesity initiatives aimed at families with potential to influence adult energy balance behaviours were also included (with child-only focused policy actions/WMS excluded from this review). Original searches aimed to capture policy actions/WMS in place from 2008-2020 relevant to NI (including those that may have been UK-wide or all-island initiatives but included NI). This time period was selected given that 2008-2009 was the year the NI Assembly Inquiry into Obesity began, and it followed the Foresight Obesity report publication (2007) which clearly illustrated the multiple and complex drivers of obesity and outlined areas for intervention (4). The updated searches captured relevant policy actions/WMS from August 2020 to early 2024.

*Exclusion:* Private services were not included (e.g., private bariatric surgery or dietetics services), however commercial WMS such as Slimming World and Weight Watchers were the exception due to their wide-use, availability, their relatively low-cost and due to a past voucher-referral scheme which enabled free access for some individuals during the inclusion period of the study. All information sources had to be in English and the most recent version of any document where available was used.

### Data extraction

In the data extraction process, relevant details of the policy actions/WMS (such as the title, document/intervention type, organisation, and year published) were recorded. Detailed information was sought where possible about each policy action/WMS from the various sources to allow for the BCW coding process and categorisation into weight management tiers. See Appendix S2 for examples of the detail captured in the data extraction sheet.

### Weight-Management Tiers

Two researchers (LM and AK) reviewed descriptions of the various interventions (policy actions/WMS) and independently coded each one, designating them to the appropriate weight-management tier, ranging from universal health promotion interventions in tier 1 to surgery in tier 4 (see criteria in figure 1). Any discrepancies were discussed, and a coding consensus was agreed upon involving a third reviewer where necessary (RON).

### Behavioural Science Mapping

A behavioural science mapping approach was adapted and applied to the policy actions/WMS identified based on the methods of Croker and colleagues (13). This entailed coding the policy actions/WMS by intervention type of the BCW Framework (intervention types described in table 1 below). Obesity risk factors (or variables) selected from the Foresight Obesity System Map were coded according to the COM-B Model.

The COM-B Model identifies the sources of behaviour that could prove fruitful targets for intervention. The constructs within the model can be subdivided as seen in table 1. The BCW provides a framework for selecting appropriate intervention types (e.g., education, training, environmental restructuring) and policy categories (e.g., fiscal measures, restrictions, guidelines, comms/marketing) based on what is known about the target behaviour/(s). Intervention types of the BCW are the approaches likely to be effective in changing one or more of the COM-B components for a behaviour change to be enacted. On the outer layer of the BCW, are seven policy categories that can support the delivery of the different intervention types (12).

### Steps in the mapping process

#### 1. Coding Policy Actions/WMS using the BCW

Each intervention was assessed for its potential to influence behaviour change through nine intervention types: education, persuasion, incentivisation, coercion, training, restriction, environmental restructuring, modelling or enablement. The Behaviour Change Wheel Book-A Guide to Designing Interventions was used as a guide (19). AK performed the coding, with a 20% sample double-coded by LM/RON for validation. Discrepancies were discussed to reach consensus, and agreement levels were calculated.

#### 2. Selecting Relevant Obesity Risk Factors

The first-tier variables (i.e. obesity risk factors) of the Foresight Map are the most strongly linked variables to the energy balance core of the map, which regulates body weight. Examples include energy density of food offerings, stress, and level of recreational activity. The first-tier variables, hereon referred to as obesity risk factors, are thought to be the most relevant targets in terms of policy levers in obesity (hence their proximity to the core of the system) and hence why they were selected for the current study. Only risk factors amenable to intervention (lifestyle change, structural policies, or medication) and those influencing adult obesity risk were included, leaving 29 factors.

Most physiological or biological factors were excluded, as they were not easily amenable to intervention (see full list and definitions of included/excluded obesity risk factors in Appendix S3).

#### 3. Coding Obesity Risk Factors with COM-B

Each obesity risk factor was coded according to the COM-B model (Capability, Opportunity, Motivation) to assess how it influences energy balance behaviours. For example, “food literacy” was coded under psychological capability, as it involves knowledge and skills to choose healthy foods.

One or more COM-B constructs could be assigned for a given obesity risk factor. Coding was done by AK, with 20% double-coded by HC. Discrepancies were discussed, and agreement was recorded.

#### 4. Systematically Mapping Policy Actions to Obesity Risk Factors

Policy actions/WMS were mapped against the selected obesity risk factors to assess how well they targeted energy balance behaviours.

The matrix of links between COM-B constructs/BCW intervention types by Michie, Atkins and West (2014) was used to specify the most appropriate intervention types for bringing about change in each COM-B construct (table 2)(19). This matrix served as a guide to compile the BCW coded policy actions/WMS and the COM-B coded obesity risk factors in a cross-reference grid. Each obesity risk factor was considered in turn, and the policy actions/WMS were cross-referenced with the BCW intervention types.

**Table 2.**
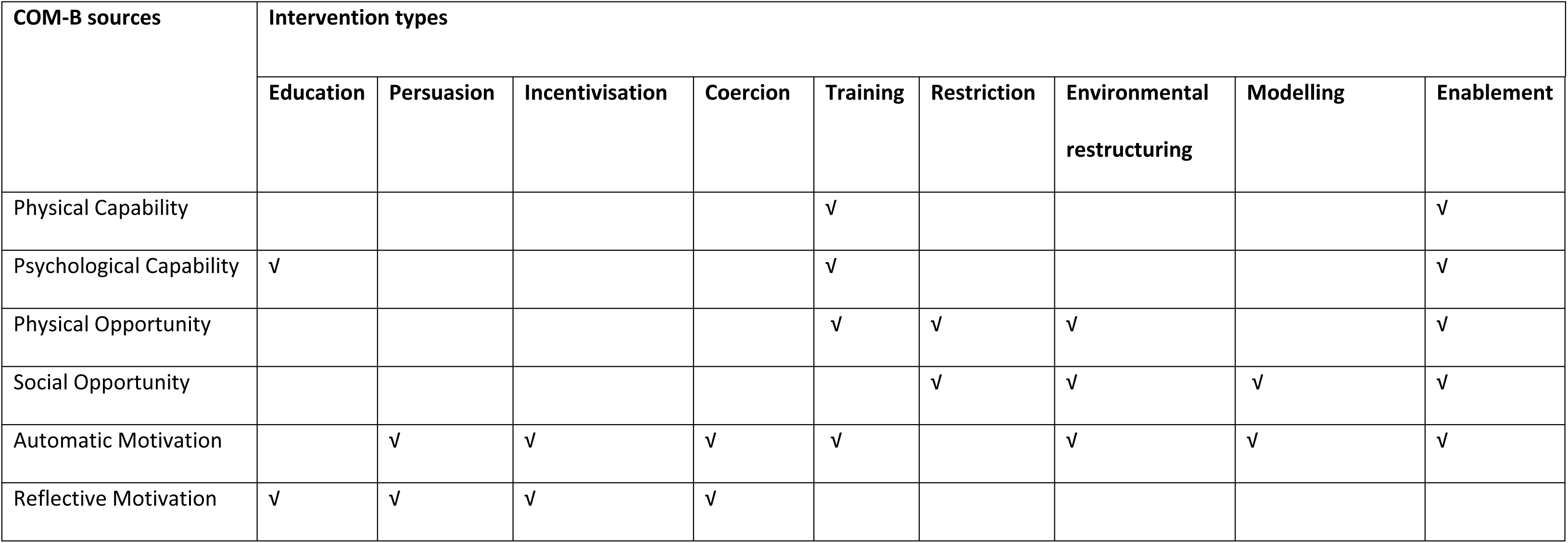
Matrix of links between COM-B and BCW intervention types (19)

| COM-B sources | Intervention types |  |  |  |  |  |  |  |  |
| --- | --- | --- | --- | --- | --- | --- | --- | --- | --- |
|  | Education | Persuasion | Incentivisation | Coercion | Training | Restriction | Environmental restructuring | Modelling | Enablement |
| Physical Capability |  |  |  |  | √ |  |  |  | √ |
| Psychological Capability | √ |  |  |  | √ |  |  |  | √ |
| Physical Opportunity |  |  |  |  | √ | √ | √ |  | √ |
| Social Opportunity |  |  |  |  |  | √ | √ | √ | √ |
| Automatic Motivation |  | √ | √ | √ | √ |  | √ | √ | √ |
| Reflective Motivation | √ | √ | √ | √ |  |  |  |  |  |

#### 5. Creating a Heat Map

The cross-referencing grid was used to generate a heat map showing where policies targeted obesity risk factors. The heat map visually displayed areas where policies were lacking or where further actions could be focused.

Grey cells on the heat map illustrated where the COM-B construct was not considered relevant for that variable according to the matrix of links between COM-B and BCW intervention types (table 2), which formed the basis of the heat map. Relevant cells on the map (i.e., whereby the specific COM-B construct was deemed relevant for that obesity risk factor) were coloured either green, red or amber as follows:

**Green**: at least one policy action/WMS in place related to that obesity risk factor, acting via that specific BCW intervention type, to target behaviour that occurred through the relevant COM-B construct.

**Red**: No policy actions/WMS identified for that risk factor via the specified intervention type/relevant COM-B construct.

**Amber**: Uncertain or unclear policy action/WMS coverage.

#### 6. Identifying Policy Gaps

The heat map revealed where policy actions/WMS were insufficient or absent, highlighting areas for further development. Policy actions/WMS were also summarised in tables specifically to show policy coverage by COM-B construct and BCW intervention type (a percentage of all relevant opportunities for policy to address obesity risk factors on the heat map, i.e. non-greyed-out cells). Percentage coverage of obesity risk factors by policy actions/WMS in both cases was considered low (less than 40%), average (40-60%), high (60-80%) or very high (>80%). Gaps where there were limited policy actions/WMS in place were seen as opportunities for new or improved policies to target obesity risk factors more effectively.

#### 7. Making Recommendations

Based on the heat map and cross-referencing results, recommendations were made to address the identified policy gaps and focus future policy actions on the most impactful obesity risk factors.

## Results

The systematic grey literature search included 12 Google Advanced Searches, three database searches, 17 searches of targeted websites and three stakeholder interviews with representatives from the Public Health Agency, SafeFood and a Health and Social Care Trust (HSCT) Dietitian, the NI equivalent of an NHS Dietitian. A total of 127 policy actions/WMS were identified as relevant, as seen in figure 3b (e.g., programmes n=45, campaigns/leaflets/online resources n=29, guidelines/standards n=20, policies/strategies/action plans n=16, services n=12 and initiatives/talks n=5). For further examples of the results found, see Appendix S2. Examples of reasons for exclusion in the first screen included anything relevant to childhood interventions only; anything not applicable to NI (e.g., England only) and any intervention existing before 2008 but not after. Removal of duplicates within the screening process and exclusion reasons are shown in figures 3a and 3b.

**Figure 3a.**
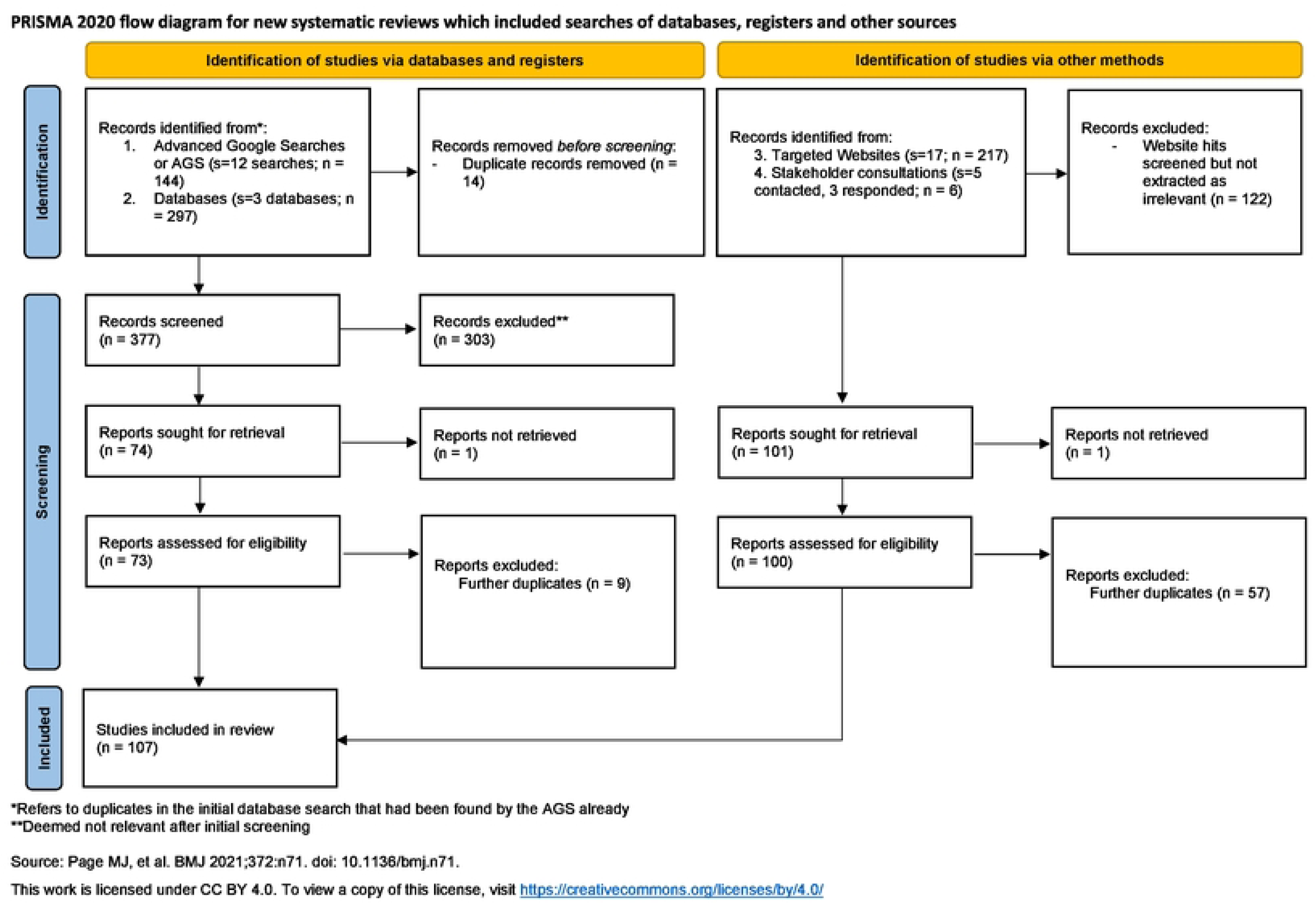
PRISMA flow diagram ORIGINAL scoping review

**Figure 3b.**
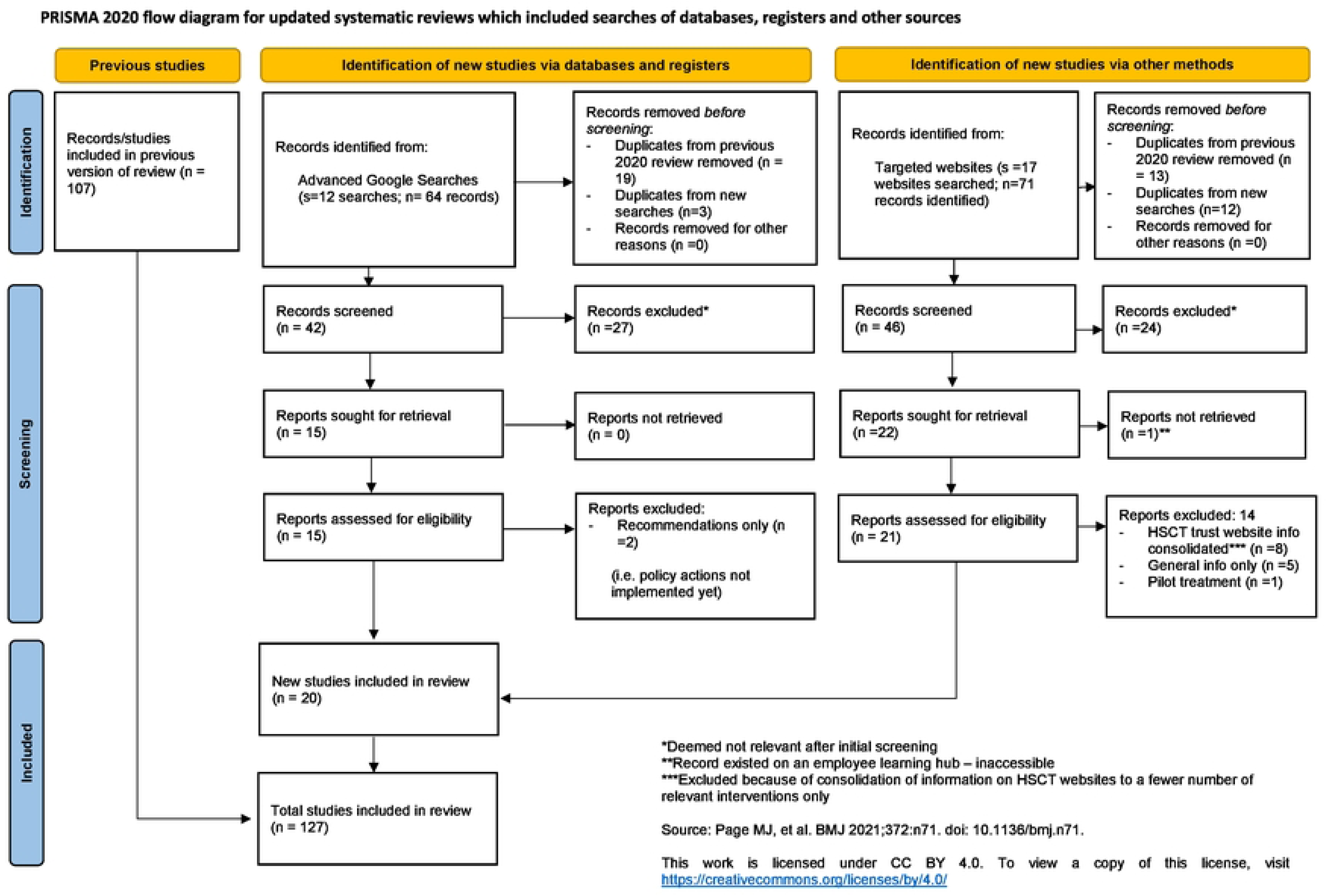
PRISMA flow diagram UPDATED scoping review

### Identifying gaps in provision by weight management tier

Full coding of search results by weight-management tier resulted in an agreement level of 91%. Most interventions for obesity in NI were classified as tier 1 interventions (72%) with a focus on general health improvement/universal healthy eating and physical activity education messaging. This included population level approaches such as the “Choose to Live Better” Campaign, SafeFood’s START Campaign and The Bicycle Strategy for Northern Ireland. A total of 21% of all interventions were classified at the tier 2 level and examples of these included programmes in community settings which offered support to individuals or groups in terms of healthy lifestyle changes (e.g., “Choose to Lose”, the “Physical Activity Referral Scheme”, and commercial evidence-based programmes such as Slimming World and Weight Watchers). Nutrition and Dietetic services were designated as combined tier 2/tier 3 services, where tier 3 is considered a more specialist obesity service (comprising 5% of all results) as they offered both these services, depending on severity of obesity.

Specialist services for weight management (tier 3) were few (2%), with only “Weigh to a Healthy Pregnancy” identified for pregnant women with obesity and “Our Hearts, Our Minds, Pan-vascular Prevention Programme” (for those at risk of or recovering from cardiovascular disease) considered to be standalone multi-disciplinary services. Publicly funded tier 4 bariatric surgery services were not found to exist, only private services were available in the NI context.

### Behavioural Science Mapping

#### Level of coding agreement

Full details of the coding files can be found in Appendix S3. The level of agreement for the policy actions/WMS coded by intervention type of the BCW was 90%, suggesting strong agreement between coders. The agreement rate for the coding of the obesity risk factors was also high at 87% (20).

Examples of how policy actions/WMS were coded by BCW intervention types are shown in Appendix S4 which is a sample of the cross-reference grid table. The most common intervention type identified was education (93% of policy actions coded for education), followed by persuasion (60%), enablement (45%) and training (36%). Some policy actions used environmental restructuring (31%), whereas incentivisation (21%) and modelling (21%) were less commonly identified. Coercion and restriction were each identified from policy actions 2% of the time (figure 4).

**Figure 4.**
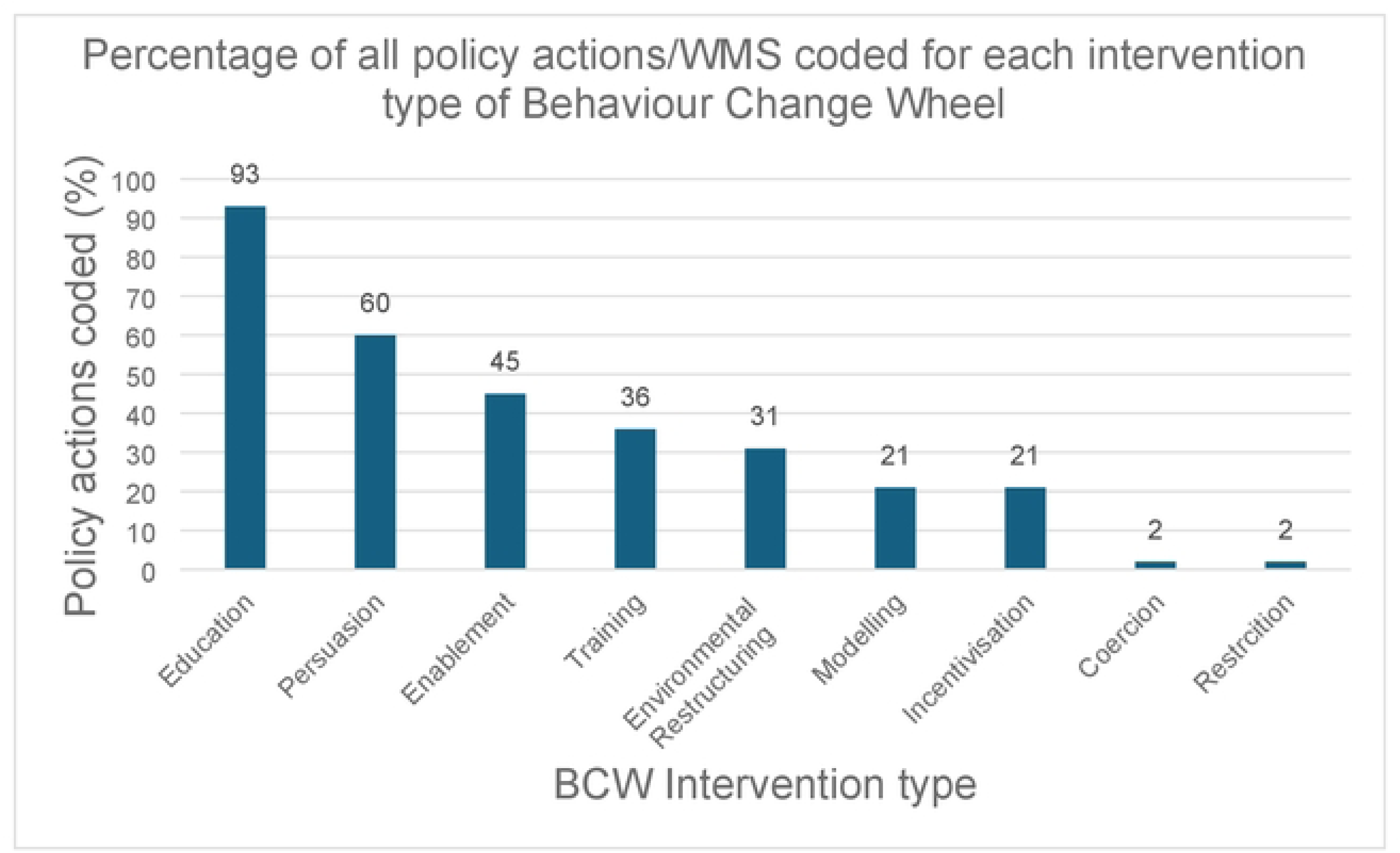
Policies/WMS by Intervention type of the BCW

#### Selecting and describing relevant obesity risk factors affecting energy balance in adults

A total of 29 obesity risk factors were selected from the first tier of the Foresight Map. The coding of the obesity risk factors resulted in a broad range of COM-B constructs being highlighted as relevant. Hence, there were a variety of drivers of adult obesity included with multiple ways for each to be potentially targeted for intervention. Figure 5 displays via a heat map how the obesity risk factors were assigned to the relevant COM-B construct(s) and then cross-referenced with appropriate intervention types according to the Matrix of links in table 2.

**Figure 5.**
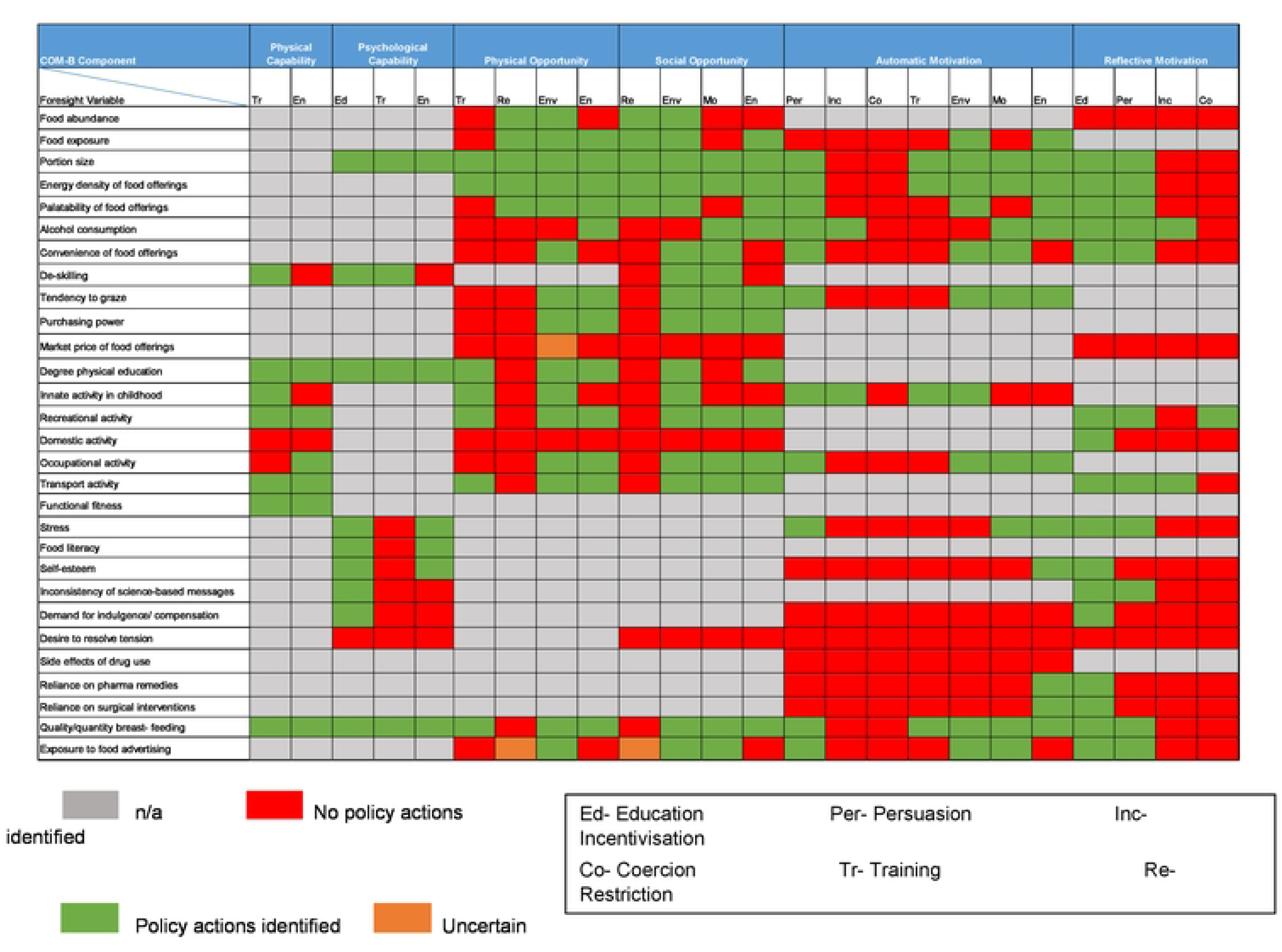
Heat Map showing the mapping of coded policy/WMS against obesity risk factors

#### Systematically overlaying the policy actions/WMS onto the selected obesity risk factors

Overall, 402 opportunities existed for obesity-related policy actions/interventions on the heat map (i.e. where an obesity risk factor mapped onto an element of COM-B and represented a behaviour change target). For example, the COM-B constructs of physical opportunity, social opportunity and automatic motivation were relevant for the obesity risk factor of ‘food exposure’ from the Foresight Map.

Overall, 48.3% (n=194) of possible opportunities had at least one policy action/WMS identified (indicated by a green coloured cell). No policy actions were identified for over half of the available opportunities (51%, n=205) or findings were unclear in a small number of cases (amber cells, n=3 or 0.7%). Tables 3 and 4 summarise the policy coverage by COM-B construct and intervention type from the BCW. Those values less than 40% were deemed ‘low coverage’ and those over 80% ‘very high coverage’ in the respective tables. Regarding percentage coverage of the COM-B constructs by available policy actions, ‘physical capability’ and ‘psychological capability’ had a coverage of 72% and 63%, respectively. Just over half of the opportunities for ‘physical’ and ‘social opportunity’ were covered by policy, and policy actions addressing both types of ‘motivation’ were fewer in number (Reflective 38%; Automatic 39%).

**Table 3.** Policies/WMS in place by COM-B construct (scored out of possible opportunities)

| COM-B Construct | Number of mapped policy actions<br>(i.e., policies/WMS) out of<br>possible opportunities | % coverage |
| --- | --- | --- |
| **Physical capability:** Physical skill, strength or stamina | 13/18 | 72 |
| **Psychological capability:** Knowledge or psychological skills, strength<br>or stamina to engage in the necessary mental processes | 19/30 | 63 |
| **Physical opportunity:** Opportunity afforded by the environment involving time, resources,<br>locations, cues, physical 'affordance' | 39/72 | 54 |
| **Social opportunity:** Opportunity afforded by interpersonal influences, social cues and cultural<br>norms that influence the way that we think about things, e.g. the words and concepts that make<br>up our language | 43/80 | 54 |
| **Reflective motivation:** Reflective processes involving plans (self-conscious intentions) and<br>evaluations (beliefs about what is good and bad) | 30/76 | 38* |
| **Automatic motivation:** Automatic processes involving emotional reactions, desires (wants and<br>needs), impulses, inhibitions, drive states and reflex responses | 48/126 | 39* |

325 \*Indicates those less than 40%, † Indicates those over 80%.

**Table 4.** Policies/WMS in place by BCW intervention type (scored out of possible opportunities)

| **BCW Intervention type** | **Number of mapped policies out of possible opportunities** | **% coverage** |
| --- | --- | --- |
| **Education:** Increasing knowledge or understanding | 25/29 | 86<sup>†</sup> |
| **Persuasion:** Using communication to induce positive or negative feelings or stimulate action | 22/37 | 59 |
| **Incentivisation:** Creating an expectation of reward | 4/37 | 11* |
| **Coercion:** Creating an expectation of punishment or cost | 1/37 | 3* |
| **Training:** Imparting skills | 22/55 | 40 |
| **Restriction:** Using rules to reduce the opportunity to engage in the target behaviour (or to increase the target behaviour by reducing the opportunity to engage in competing behaviours) | 10/38 | 26* |
| **Environmental restructuring:** Changing the physical or social context | 41/56 | 73 |
| **Modelling:** Providing an example for people to aspire to or imitate | 21/38 | 55 |
| **Enablement:** Increasing means/reducing barriers to increase capability (beyond education and training) or opportunity (beyond environmental restructuring) | 48/75 | 64 |

\*Indicates those less than 40%, † Indicates those over 80%.

Within the policy actions/WMS addressing obesity risk factors identified in the mapping review, a wide range of BCW intervention types were noted (table 4). The coverage of available opportunities for policy actions by intervention type, mapped against the obesity risk factors, was highest for education (86%). Most policy actions/WMS identified in the searches were coded as using education, either alone or combined with other intervention types, such as persuasion or enablement. The intervention types of environmental restructuring and enablement had moderate coverage, with many opportunities for these intervention types having at least some policy actions/WMS in place (73% and 64% respectively). Policy coverage ranged from 40-59% for persuasion, training and modelling. Coverage of possibilities for restriction was 26%, which related mainly to one policy action (Minimum Nutritional Standards for Health and Social Care).

Incentivisation (11%) and coercion (3%) were among the least covered by available policy actions/WMS.

#### Coverage of specific obesity risk factors by policy actions/WMS

##### Food consumption

Obesity risk factors most often addressed by policy actions/WMS related to food and nutrition were ‘portion size’ and ‘energy density of food,’ followed by ‘quality/quantity of breastfeeding’ and ‘food literacy’. Policy actions/WMS that addressed these food-related obesity risk factors included campaigns, initiatives, and community programmes. An example to illustrate this in the case of ‘energy density of food’, were campaigns to increase awareness of the calories in food such as ‘Choose to Live Better’ and the ‘Eating Well, Choosing Better’ initiative to encourage food companies to reformulate or offer less energy dense alternatives (see Appendix S2 for details of these initiatives). Food-related risk factors with limited coverage included ‘Market price of food’ and to a degree ‘Exposure to food advertising’.

##### Physical activity

There were several policy actions/WMS for ‘level of recreational activity’, ‘transport activity’, ‘functional fitness’, and ‘degree of physical education’ (with over 70% coverage). Examples included Active Travel programmes and National Cycle Network training.

##### Psychology/Physiology

There were a lack of policy actions/WMS to address psychological and impulsive/emotion-driven variables such as ‘demand for indulgence/compensation’, ‘desire to resolve tension’ and ‘self-esteem’. Many of the other physiological/biological variables were excluded from the present review due to lack of factor modifiability.

## Discussion

This systematic scoping review identified and mapped a range of policy actions/WMS relating to adult obesity prevention and management in one part of the UK (NI) from 2008-2024. Categorisation of policy actions/WMS (n=111 excluding NICE Guidelines) by weight-management tiers illustrated an over-reliance on interventions focused on general health promotion (tier 1). Coding policy actions/WMS using the BCW indicated that a wide range of intervention types were employed, although reliance on educational and persuasive campaigns (information-giving) predominated.

Scope exists for significant policy development utilising a wider variety of behaviour change methods focusing on environmental restructuring, incentivisation, modelling, restriction and enablement.

Policy actions/WMS were mapped against obesity risk factors identified from the Foresight Obesity System Map to illustrate how comprehensively existing policy actions/WMS targeted these risk factors (4). Despite policy actions/WMS targeting a wide variety of obesity risk factors, there were significant gaps in relation to several important drivers such as ‘exposure to food advertising’, ‘demand for indulgence/compensation’, ‘desire to resolve tension’, ‘self-esteem’ and ‘side effects of drug use’. Findings also indicated a lack of policy actions/WMS at the COM-B level of automatic and reflective motivation, for example interventions to reduce exposure to unhealthy food environments or equipping individuals with strategies to make healthier choices in response to the food environment.

This systematic behavioural science mapping approach, similar to one used in previous obesity research (13) identified policy actions/WMS for less than half (48.3%) of all potential opportunities for addressing energy balance drivers, displayed via a heat map. Significant scope for a wider breadth of intervention approaches via policy actions/WMS was noted.

### Weight-management tiers

The majority of policy measures identified within the “A Fitter Future For All” strategic framework for preventing and addressing overweight and obesity relied heavily on individual responsibility to make behavioural changes, largely focussing on health promotion/education. This approach aligns with the UK’s historical trend of successive governments adopting less interventionist policies over the past 30 years, with ineffective implementation and poor evaluation also hampering progress according to a recent analysis of UK obesity policies and strategies (3) . While a small number of initiatives in NI, such as the “Minimum Nutritional Standards for Health and Social Care,” aim to alter the food environment, evidence suggests that multicomponent workplace interventions (e.g., involving healthier food provision in cafeterias, food vending machines) can induce positive health-related behaviour changes (21), therefore implementing more of these should be considered (21).

Nutritional standards and the introduction of the Florence weight management service (via an app in 2021) in HSCT settings is a positive step forward in terms of workplace interventions for those working in healthcare.

Community WMS in tier 2 (e.g., Choose to Lose) have had some short-term success in NI, but they have not been commissioned long enough to be evaluated medium-long term. There are some examples of more (traditionally considered) tier 2 programmes being rolled out recently, as evidenced by the updated search in 2024. Examples include Safefood’s 12-week weight loss programme ‘A Healthy Weight For You’ and a Momenta Weight Management programme available in one local HSCT. Wider commissioning of these services is needed and an absence of ongoing support for individuals with obesity is a concern. Systematic review evidence suggests that weight loss interventions (with follow up of more than one year) may reduce premature all-cause mortality in those with obesity, however more evidence on the longer-term weight loss/maintenance outcomes of weight-management programmes is needed (22, 23).

No published studies or evaluations of tier 2/3 adult WMS in NI were found in searches. The roll out of an evidence-based behaviour change programme across NI (the Diabetes Prevention Programme introduced in 2019) is a positive development. However, urgent action is needed to develop a community weight-management offering available to all adults in need of weight-management support in NI, beyond those with pre-diabetes. A recent systematic review has shown that referral to a commercial weight management programme delivered in the community is cost-effective and benefits public health in the long term, yielding higher quality-adjusted life-years (QALY) than brief intervention, thus reinforcing existing evidence supporting behaviour change programmes for obesity (24).

The positive impact of tier 3 WMS on clinical outcomes in other parts of the UK has been demonstrated previously (25). However, no established NHS tier 3 obesity service in NI for those with complex obesity represents a significant gap in service provision as per NICE guidelines (CG189) (26). Indeed, the lack of prescription of (or absence of) weight loss medications as part of NHS WMS was notable in this region of the UK (NI). Whilst some of these (e.g., GLP-1 agonists) have been NHS-prescribed and in use privately for obesity throughout the UK for some time now, NI lags behind the rest of the UK in terms of rolling out new advances in obesity medications, with only Orlistat approved for use in NI HSC settings, usually in primary care. Since 2021, the US Food and Drug Administration (FDA) have approved the use of six medications for weight management in adults with a BMI ≥ 30kg/m² or ≥27kg/m² with comorbidities. These are Orlistat, Phentermine plus Topiramate, Naltrexone plus Bupropion, Liraglutide, Semaglutide and more recently Tirzepatide (as brand name Zepbound for weight loss) (Yanovski and Yanovski, 2021). Liraglutide and Semaglutide were only approved by the FDA in 2015, an indication of how recent these advances in pharmacotherapy for obesity are. In February 2022, NICE recommended Semaglutide (Wegovy) for obesity in the UK, with guidance published in 2023 (27). Liraglutide (Saxenda) and Semaglutide are prescribed by specialist WMS in England, Scotland, and Wales, with Semaglutide showing greater weight loss than Liraglutide (28). Additionally, NICE approved Mounjaro (Tirzepatide) for use in primary care in the UK last year, with roll out due in Spring 2025 (29). However, in Northern Ireland, none of these GLP-1 drugs are prescribed through the HSC and they are not listed on the Northern Ireland Medicines and Healthcare products Regulatory Agency (MHRA) Authorised Route (NIMAR), limiting weight management options and creating inequalities in obesity care compared to the rest of the UK.

This study found no availability of HSC-funded tier 4 services in NI, again highlighting inadequate care for those living with obesity. According to NICE Guidelines (NG246 now replacing CG189), bariatric surgery is recommended as a treatment for individuals with recent-onset Type 2 Diabetes (within 10 years onset) with expedited assessment for those with a BMI of 35kg/m² or above (30). Long been seen as a last resort, there is now no requirement to fully utilise all non-surgical options before being considered for surgery (31). Current NI levels of adult obesity (28%) and effectiveness data on bariatric surgery suggest a need for this treatment modality to be available (32, 33). The lack of provision of bariatric surgery often forces those ‘would-be’ eligible candidates for surgery in the UK/Ireland to either pay privately (at home or abroad) or go without if they cannot afford the expensive procedure and after-care (typically around £12,000) (34, 35). Seeking treatment abroad in particular, can have negative implications for access to appropriate post-operative care and support needed to adjust to life after surgery (36).

Adopting an integrated approach to weight-management comprising a simplified, more flexible pathway has been recommended in the UK where patients can move more freely between the levels of treatment available (8). A system with one tier for prevention and another tier for management was proposed, with NICE considering tiers 3 and 4 together as specialist multidisciplinary obesity services in their most recent update (30). Based on the results of this review, obesity management is an area that needs significant development and investment in NI; given the over-reliance on prevention-focused obesity policy/WMS to date, which typically come under the tier 1 remit of the traditionally used tiered system. Any new developments in progress to improve the support offered to those living with obesity in NI, should take into consideration the evidence-base and the changing landscape of obesity services in the UK as a whole (i.e., whether the current tiered system for weight management is fit for purpose).

### Behavioural science mapping

This scoping review highlighted examples of environmental restructuring through infrastructural changes, such as the recent investment in Greenways and the promotion of healthier, cleaner transportation methods. These changes may have positively influenced physical activity rates, as similar initiatives have increased activity levels in other regions(37), whether this translates into an impact on reduction of population obesity rates remains to be established. A recent natural experiment by Hunter and colleagues, 2021 evaluated the physical activity, health, wellbeing, social and environmental effects of a new urban greenway in NI (PARC study) (38). The study did not find significant population-level effects on physical activity or mental wellbeing, however the need for longer term evaluation of impact was stressed (38). The findings from this study indicate scope to develop more interventions using different forms of environmental restructuring in NI.

The findings of this review align with themes found in obesity policy literature from high-income countries in particular, and specifically within the UK (39). These themes highlight a focus on education and interventions that rely on individuals making ’good choices,’ with less emphasis on structural changes or measures that limit individual choice. However, structural changes, which bypass or limit self-conscious decision-making, may be more effective in supporting behaviour change, as they reduce reliance on reflective motivation (3). Structural policies also have the added benefit of being more likely to reduce health inequalities compared to those focusing on individual agency (40). One example is the UK Soft Drinks Industry Levy, a recent analysis of which has shown the greatest impact on sugar consumption among lower-income households with children (41).

A recent toolkit compares over 30 obesity and health-focused policies for the UK, assessing evidence quality, impact on obesity, and cost to government to aid policymakers in planning effective interventions (42). The report suggests that a policy package combining tax, regulation, and treatment would have the greatest health impact at the lowest cost, though a prevention and treatment package could achieve similar impact (at similar cost) without the political opposition to food taxes. This package includes mandatory health targets for retailers, restrictions on HFSS food promotions, and expanded access to pharmacological weight loss treatments. Uptake and implementation of such policies remain uncertain in the current UK context. Growing support for system-level interventions versus individual-focussed measures (described as “s-frame” and “i-frame” interventions by Chater and Loewenstein) is evident, albeit both types must work in tandem for optimal outcomes (43).

WSAs are considered particularly important in terms of future direction in reducing obesity levels (7). A key challenge with WSAs for obesity is evaluation, with many methods deemed to be in their infancy (44, 45). However, examples of evaluations and good practice elsewhere could be adopted such as the Amsterdam Healthy Weight Approach, where the prevalence of overweight/obesity among 2-18-year-olds has decreased from 21% in 2012 to 18.7% in 2017, an early indication of success in a WSA for obesity (6). Learnings from the current mapping review should be considered in future approaches to obesity in NI, with care taken to effectively incorporate obesity treatment and management options into the WSA given the significant proportions of the population living with excess weight (28% with obesity) (32). Ultimately, effective WSAs should focus not only on prevention but also on ensuring access to effective treatments (46).

The COM-B Framework explains how health behaviour change can be considered in the context of three interacting elements: improving people’s capabilities (their knowledge or skills), their opportunities (physical or social) and their motivations. In terms of how BCW intervention types mapped against the identified COM-B constructs, education was the most frequently coded intervention under psychological capability (relying on increasing people’s knowledge), which is unsurprising considering the percentage of policy actions/WMS in the overall mapping that used education. The majority of policy actions/WMS fell into the tier 1 category of weight-management which is in line with the finding of educational measures and ‘information-giving’ being common (as is the norm with universal health promotion messages). Training was under-represented for psychological capability. Targeting food literacy for example could be an effective training intervention in NI (e.g., training individuals to read and properly understand food labelling and nutritional information).

The coverage of physical and psychological capability by mapped policy actions/WMS in this study differed slightly from the 2020 study, with coverage at 72% vs 93% for physical capability, and 63% vs 54% for psychological capability, respectively (13). Most physical capability-related actions in both studies focused on physical activity, where training (e.g., skills sports/exercise) and enablement (e.g., providing bicycles for transport/leisure) were key intervention types. Gaining cooking skills was categorized as training. If effective, cookery programs in NI could improve cooking skills, potentially benefiting energy balance behaviours, as reduced home cooking has been linked to rising obesity levels over time (47).

There were many opportunities for policy actions/WMS relevant to both types of COM-B opportunity constructs and they spanned a wide range of obesity risk factors (related to diet, physical activity and economy). Opportunities for policy actions related to these constructs received moderate coverage, with over half having numerous policy actions/WMS identified. The intervention type of restriction was not coded for many opportunities and where it was, the Minimum Nutritional Standards for Health and Social Care guidelines document was one of the only policies/WMS coded for this intervention type. There was scope to increase both physical/social opportunities related to dietary factors such as alcohol consumption and convenience of food offerings, such as interventions to make more healthy convenience food options available.

Both types of motivation were the least covered constructs, despite being relevant for most obesity risk factors. Automatic motivation had the most potential intervention types (seven), indicating numerous possible approaches, yet many obesity risk factors (e.g., self-esteem, demand for indulgence/compensation, desire to resolve tension, side effects of drug-use, reliance on pharma/surgical remedies and exposure to food advertising) lacked policies targeting automatic motivation. There is potential to develop policies using training, incentivisation, coercion, modelling, and environmental restructuring to address automatic processes. For reflective motivation, incentivisation and coercion also offer potential, with examples like food vouchers to purchase healthy foods in the supermarket, increasing an individual’s motivation to buy healthier foods.

Exposure to food advertising for adults was underrepresented in policy actions, despite being a significant obesity risk factor, particularly for children but also impacting adults’ eating motivations (48, 49). Often governments prioritise children’s exposure to advertising which is the case with current UK-wide plans for TV/online restrictions, however these plans have experienced multiple delays and are yet to be implemented. This gap presents an opportunity for effective interventions targeting both adults and children in NI, influencing motivation. Additionally, this study highlights that restriction is an underutilised intervention type generally, despite evidence suggesting benefits of limiting food advertising to children elsewhere (50).

Capability had higher coverage than opportunity in this mapping review, again suggesting a reliance on individual capabilities and responsibility for behaviour change. This contrasts with the relatively low availability of policy actions/WMS to enhance the physical and social opportunity of individuals to change behaviour. Together with limited policy actions/WMS acting at the level of motivation, this reinforces what has been discussed in this review and elsewhere about the over-reliance on individual agency in obesity prevention and management (3).

### Strengths and limitations

This systematic scoping review mapped obesity policy actions and WMS in Northern Ireland, using an innovative behavioural science informed approach. It offers valuable insights into the available services since 2008 and provides a strong foundation to develop future policies/services, allowing further exploration of gaps in provision where they were found. The study covered national, regional, and local policies, and included small-scale interventions, providing a comprehensive overview of the policy/service landscape (a strength of the study). This approach not only contributes to local policy in NI but also has broader applicability for other regions. The contribution of this study to literature and policymaking over and above the local application in NI is of note also, with potential for the methodology to be applied more broadly elsewhere.

The grey literature search strategy proved a useful tool and was a strength of the methodology however it was subject to limitations. Engaging with key healthcare stakeholders who were dealing with workload backlogs from the Covid-19 pandemic proved challenging, making it difficult to verify availability of more specialist services across all regions of NI. Additionally, Google Advanced Searches had a built-in relevancy ranking, which might limit results, though saturation seemed to occur within the first 10 pages of searches. The study did not evaluate the implementation or effectiveness of the policies or WMS, a crucial area for understanding why current obesity interventions in NI are not achieving the desired outcomes. Finally, the Foresight Obesity System Map (2007) may not reflect newer obesity risk factors, such as sleep or the influence of social media, which are now recognised in more recent research (51).

## Conclusion

Systematically identifying and mapping obesity-relevant policy actions/WMS across the country of NI highlighted gaps in provision and opportunities for development both from the perspective of tiered weight-management provision and from a behavioural science perspective. Most interventions focused on general health promotion and limited specialist services existed to effectively manage obesity for the almost one million adults currently living with overweight or obesity in NI. A more flexible approach to weight-management such as having separate prevention and management pathways, as suggested by Hazlehurst and colleagues recently, may also be of benefit. Behavioural science mapping using the BCW Framework and the COM-B Model indicated that most interventions were education-based with scope for increasing the number of policy actions/WMS using other intervention types, and overall, a lack of interventions to support and enable public motivation to enact healthy behavioural changes must be addressed.

## Data Availability

All relevant data are within the manuscript and its Supporting Information files.

## Note on funding

The main author of this work (AK) was funded by a Department for the Economy (DfE) Northern Ireland studentship. No other funding was received. The funder had no role in the conduct, analysis or interpretation of the results.

## Acknowledgements

The authors would like to thank the stakeholders involved in the consultations that were conducted as part of the scoping exercise of obesity policies in NI. Special thanks also to Ms Sara Lucas for updating the grey literature searches.

## Supporting information captions

S1 Appendix-Grey literature search details

S2 Appendix-Data Extraction sheet

S3 Appendix-Coding file

S4 Appendix-Cross-reference grid sample

S5 PRISMA-ScR Checklist

## Notes

### Competing Interest Statement

The authors have declared no competing interest.

### Funding Statement

Yes

### Author Declarations

Queens University Faculty Research Ethics Committee

## References

1. Seidell JC, Halberstadt J. The Global Burden of Obesity and the Challenges of Prevention. Annals of Nutrition and Metabolism. 2015;66(2):7–12.

2. Safaei M, Sundararajan EA, Driss M, Boulila W, Shapi’i A. A systematic literature review on obesity: Understanding the causes & consequences of obesity and reviewing various machine learning approaches used to predict obesity. Computers in Biology and Medicine. 2021;136:104754.

3. Theis DRZ, White M. Is Obesity Policy in England Fit for Purpose? Analysis of Government Strategies and Policies, 1992–2020. The Milbank Quarterly. 2021;99(1):126–70.

4. Butland B, Jebb S, Kopelman P, McPherson K, Thomas S, Mardell J, et al. Foresight. Tackling obesities: future choices. Project report. Foresight Tackling obesities: future choices Project report. 2007.

5. Adams J, Mytton O, White M, Monsivais P. Why Are Some Population Interventions for Diet and Obesity More Equitable and Effective Than Others? The Role of Individual Agency. PLOS Medicine. 2016;13(4):e1001990.

6. Sawyer A, den Hertog K, Verhoeff AP, Busch V, Stronks K. Developing the logic framework underpinning a whole-systems approach to childhood overweight and obesity prevention: Amsterdam Healthy Weight Approach. Obes Sci Pract. 2021;7(5):591–605.

7. Backholer K, Beauchamp A, Ball K, Turrell G, Martin J, Woods J, et al. A framework for evaluating the impact of obesity prevention strategies on socioeconomic inequalities in weight. Am J Public Health. 2014;104(10):e43–50.

8. Hazlehurst JM, Logue J, Parretti HM, Abbott S, Brown A, Pournaras DJ, et al. Developing Integrated Clinical Pathways for the Management of Clinically Severe Adult Obesity: a Critique of NHS England Policy. Curr Obes Rep. 2020;9(4):530–43.

9. Coulton V, Dodhia SK, Ells L, Blackshaw J, Tedstone A, editors. National mapping of weight management services: provision of tier 2 and tier 3 services in England 2015.

10. Read S, Logue J. Variations in weight management services in Scotland: a national survey of weight management provision. J Public Health (Oxf). 2016;38(3):e325–e35.

11. Sharman K, Nobles J. Bridging the gap: SHINE – a Tier 3 service for severely obese children and young people. The British Journal of Obesity. 2015;1:158–63.

12. Michie S, van Stralen MM, West R. The behaviour change wheel: A new method for characterising and designing behaviour change interventions. Implementation Science. 2011;6(1):42.

13. Croker H, Russell SJ, Gireesh A, Bonham A, Hawkes C, Bedford H, et al. Obesity prevention in the early years: A mapping study of national policies in England from a behavioural science perspective. PLOS ONE. 2020;15(9):e0239402.

14. Allender S, Owen B, Kuhlberg J, Lowe J, Nagorcka-Smith P, Whelan J, et al. A Community Based Systems Diagram of Obesity Causes. PLOS ONE. 2015;10(7):e0129683.

15. Savona N, Macauley T, Aguiar A, Banik A, Boberska M, Brock J, et al. Identifying the views of adolescents in five European countries on the drivers of obesity using group model building. Eur J Public Health. 2021.

16. Esdaile E, Thow AM, Gill T, Sacks G, Golley R, Love P, et al. National policies to prevent obesity in early childhood: Using policy mapping to compare policy lessons for Australia with six developed countries. Obesity Reviews. 2019;20(11):1542–56.

17. Munn Z, Peters MDJ, Stern C, Tufanaru C, McArthur A, Aromataris E. Systematic review or scoping review? Guidance for authors when choosing between a systematic or scoping review approach. BMC Medical Research Methodology. 2018;18(1):143.

18. Godin K, Stapleton J, Kirkpatrick SI, Hanning RM, Leatherdale ST. Applying systematic review search methods to the grey literature: a case study examining guidelines for school-based breakfast programs in Canada. Systematic Reviews. 2015;4(1):138.

19. Michie S, Atkins L, West R. The behaviour change wheel. A guide to designing interventions 1st ed Great Britain: Silverback Publishing. 2014:1003–10.

20. McHugh ML. Interrater reliability: the kappa statistic. Biochemia Medica. 2012:276–82.

21. Naicker A, Shrestha A, Joshi C, Willett W, Spiegelman D. Workplace cafeteria and other multicomponent interventions to promote healthy eating among adults: A systematic review. Preventive Medicine Reports. 2021;22:101333.

22. Ma C, Avenell A, Bolland M, Hudson J, Stewart F, Robertson C, et al. Effects of weight loss interventions for adults who are obese on mortality, cardiovascular disease, and cancer: systematic review and meta-analysis. BMJ. 2017;359:j4849.

23. Varkevisser RDM, Van Stralen MM, Kroeze W, Ket JCF, Steenhuis IHM. Determinants of weight loss maintenance: a systematic review. Obesity Reviews. 2019;20(2):171–211.

24. Ahern AL, Breeze P, Fusco F, Sharp SJ, Islam N, Wheeler GM, et al. Effectiveness and cost-effectiveness of referral to a commercial open group behavioural weight management programme in adults with overweight and obesity: 5-year follow-up of the WRAP randomised controlled trial. The Lancet Public Health. 2022;7(10):e866–e75.

25. Brown TJ, O’Malley C, Blackshaw J, Coulton V, Tedstone A, Summerbell C, et al. Exploring the evidence base for Tier 3 weight management interventions for adults: a systematic review. Clinical Obesity. 2017;7(5):260–72.

26. Welbourn R, Dixon J, Barth JH, Finer N, Hughes CA, Le Roux CW, et al. NICE-accredited commissioning guidance for weight assessment and management clinics: a model for a specialist multidisciplinary team approach for people with severe obesity. Obesity surgery. 2016;26:649–59.

27. NICE. Semaglutide for managing overweight and obesity TA875 2023. Available from: https://www.nice.org.uk/guidance/ta875.

28. Rubino DM, Greenway FL, Khalid U, O’Neil PM, Rosenstock J, Sørrig R, et al. Effect of Weekly Subcutaneous Semaglutide vs Daily Liraglutide on Body Weight in Adults With Overweight or Obesity Without Diabetes: The STEP 8 Randomized Clinical Trial. Jama. 2022;327(2):138–50.

29. NICE. Tirzepatide for managing overweight and obesity TA1026 2024. Available from: https://www.nice.org.uk/guidance/ta1026.

30. NICE. Overweight and obesity management NG246 2025. Available from: https://www.nice.org.uk/guidance/ng246.

31. O’Hara H, Miras AD. Shift the paradigm to shift the weight: obesity care in the community. Br J Gen Pract. 2024;74(743):275–8.

32. Information Analysis Directorate DoHDNI. Health Survey (NI): First Results 2023/24. 2024.

33. Xia Q, Campbell JA, Ahmad H, Si L, Graaff B, Palmer AJ. Bariatric surgery is a cost-saving treatment for obesity—A comprehensive meta-analysis and updated systematic review of health economic evaluations of bariatric surgery. Obesity Reviews. 2020;21(1).

34. Healy P, Clarke C, Reynolds I, Arumugasamy M, McNamara D. Complications of bariatric surgery – What the general surgeon needs to know. The Surgeon. 2016;14(2):91–8.

35. Doble B, Wordsworth S, Rogers CA, Welbourn R, Byrne J, Blazeby JM, et al. What Are the Real Procedural Costs of Bariatric Surgery? A Systematic Literature Review of Published Cost Analyses. Obesity Surgery. 2017;27(8):2179–92.

36. McGrice M, Don Paul K. Interventions to improve long-term weight loss in patients following bariatric surgery: challenges and solutions. Diabetes, Metabolic Syndrome and Obesity: Targets and Therapy. 2015:263.

37. Stappers NEH, Van Kann DHH, Ettema D, De Vries NK, Kremers SPJ. The effect of infrastructural changes in the built environment on physical activity, active transportation and sedentary behavior - A systematic review. Health Place. 2018;53:135–49.

38. Hunter RF, Adlakha D, Cardwell C, Cupples ME, Donnelly M, Ellis G, et al. Investigating the physical activity, health, wellbeing, social and environmental effects of a new urban greenway: a natural experiment (the PARC study). International Journal of Behavioral Nutrition and Physical Activity. 2021;18(1).

39. Ulijaszek SJ, McLennan AK. Framing obesity in UK policy from the Blair years, 1997-2015: the persistence of individualistic approaches despite overwhelming evidence of societal and economic factors, and the need for collective responsibility. Obes Rev. 2016;17(5):397–411.

40. Adams J. Addressing socioeconomic inequalities in obesity: Democratising access to resources for achieving and maintaining a healthy weight. PLOS Medicine. 2020;17(7):e1003243.

41. Pell D, Mytton O, Penney TL, Briggs A, Cummins S, Penn-Jones C, et al. Changes in soft drinks purchased by British households associated with the UK soft drinks industry levy: controlled interrupted time series analysis. BMJ. 2021:n254.

42. Nesta Bb. Blueprint for halving obesity: A toolkit for designing solutions to reduce obesity. 2024.

43. Chater N, Loewenstein G. The i-frame and the s-frame: How focusing on individual-level solutions has led behavioral public policy astray. Behavioral and Brain Sciences. 2023;46:e147.

44. Bagnall A-M, Radley D, Jones R, Gately P, Nobles J, Van Dijk M, et al. Whole systems approaches to obesity and other complex public health challenges: a systematic review. BMC Public Health. 2019;19(1):8.

45. Karacabeyli D, Allender S, Pinkney S, Amed S. Evaluation of complex community-based childhood obesity prevention interventions. Obesity Reviews. 2018;19(8):1080–92.

46. Research NIfH. How can local authorities reduce obesity? Insights from NIHR research. 2022.

47. Seguin RA, Aggarwal A, Vermeylen F, Drewnowski A. Consumption Frequency of Foods Away from Home Linked with Higher Body Mass Index and Lower Fruit and Vegetable Intake among Adults: A Cross-Sectional Study. Journal of Environmental and Public Health. 2016;2016:1–12.

48. Smith R, Kelly B, Yeatman H, Boyland E. Food Marketing Influences Children’s Attitudes, Preferences and Consumption: A Systematic Critical Review. Nutrients. 2019;11(4):875.

49. Boyland EJ, Burgon RH, Hardman CA. Reactivity to television food commercials in overweight and lean adults: Physiological, cognitive and behavioural responses. Physiology & Behavior. 2017;177:182–8.

50. Coleman PC, Hanson P, van Rens T, Oyebode O. A rapid review of the evidence for children’s TV and online advertisement restrictions to fight obesity. Preventive Medicine Reports. 2022;26:101717.

51. Ogilvie RP, Patel SR. The epidemiology of sleep and obesity. Sleep Health. 2017;3(5):383–8.

